# Hepatic and abdominal adiposity in type 2 diabetes as assessed with machine learning on CT scans

**DOI:** 10.64898/2025.12.16.25342409

**Authors:** Richard H. Tran, Pavan Raghupathy, Mohamad Hazim, Elizabeth Thompson, Sophia Swago, Abhijit Bhattaru, Matthew MacLean, Jeffrey T. Duda, James Gee, Charles Kahn, Daniel J. Rader, Arijitt Borthakur, Walter R. Witschey, Hersh Sagreiya

**Affiliations:** Department of Radiology, Perelman School of Medicine, University of Pennsylvania, Philadelphia, PA, USA; University of Massachusetts Chan Medical School, Worcester, MA; Department of Genetics, Perelman School of Medicine, University of Pennsylvania, Philadelphia, PA, USA; Leonard Davis Institute of Health Economics, University of Pennsylvania, Philadelphia, PA, USA

**Keywords:** type 2 diabetes, hepatic steatosis, computed tomography, abdominal adiposity, visceral adipose tissue, subcutaneous adipose tissue, machine learning, artificial intelligence

## Abstract

**Aims:** The distribution of abdominal adipose depots and their mechanistic links to type 2 diabetes remain incompletely understood. This study elucidated the relationship between type 2 diabetes presence and quantitative abdominal imaging traits, including hepatic steatosis, liver and spleen size, and adipose distribution, using unenhanced computed tomography (CT) scans from a large-scale, racially diverse, disease-focused medical biobank.

**Materials and Methods:** Deep learning algorithms were applied to abdominal CT scans to automatically quantify image-derived phenotypes, including spleen-hepatic attenuation difference (SHAD) for hepatic steatosis, liver and spleen volumes (LV and SV, respectively), visceral and subcutaneous adipose tissue (VAT and SAT, respectively), and visceral-to-subcutaneous fat ratio (VSR).

**Results:** Diabetic individuals demonstrated a greater degree of hepatic steatosis and central adiposity than those without diabetes. Liver attenuation was lower (47.6 vs. 52.4 Hounsfield units (HU); lower values indicate greater steatosis), SHAD was higher (−5.41 vs. −8.41 HU; more positive values indicate greater steatosis), and steatosis prevalence increased (38.4% vs. 21.4%) (all p<2.2×10⁻¹⁶). VSR was also elevated (0.64 vs. 0.54, p=5.86×10⁻¹³). These trends remained significant after stratification by sex. Multivariate analyses revealed independent associations of diabetes with SHAD (OR 1.04), LV (OR 1.59), SV (OR 3.95), VAT (OR 1.23), SAT (OR 1.05), and VSR (OR 2.27), after adjusting for age, sex, race, and BMI.

**Conclusions:** Hepatic steatosis, hepatomegaly, and visceral adiposity on CT imaging are predictive of type 2 diabetes presence. Notably, VSR showed a stronger association with diabetes than BMI, underscoring how body-fat distribution, rather than overall adiposity, more accurately reflects metabolic disease risk.

## Introduction

Diabetes mellitus currently impacts more than 500 million people worldwide and is estimated to increase to 780 million people by 2045, with 90% classified as type 2 diabetes mellitus [1, 2]. Type 2 diabetes is diagnosed with serum hemoglobin A1c (HbA1c) > 6.5% [3]. However, challenges in healthcare access, underutilization of primary care resources, and insufficient diabetes screening programs delay disease diagnosis, leading to missed opportunities for early intervention that can prevent serious associated cardiometabolic diseases [3–5]. Given these challenges, there is a clinical need to utilize different methods, such as imaging, as opportunistic screening tools, whereby information obtained during routine or unrelated clinical assessments can be leveraged for type 2 diabetes presence and its associated risk factors.

With over 85 million computed tomography (CT) scans performed in the United States yearly, there is the opportunity to harness such data to identify patients with type 2 diabetes [6]. Recent developments in machine learning, especially within radiology, allow for rapid analysis of substantial amounts of data [7]. Recent studies have applied machine learning to assess for stroke, osteoporosis, and sarcopenia on CT scans [8–10]. Machine learning has also been utilized to diagnose and follow the progression of metabolic dysfunction-associated liver disease (MASLD) on CT; this disease reflects excess fat buildup in the liver, commonly known as hepatic steatosis (HS), which can progress to cirrhosis and hepatocellular carcinoma [11–13]. As MASLD is marked by hepatic steatosis, non-contrast CT provides an alternative to measure hepatic steatosis through the spleen-hepatic attenuation difference (SHAD). Although liver biopsy is considered the gold standard, it is invasive and can be affected by sampling error; other diagnostic methods such as magnetic resonance imaging are expensive, and traditional ultrasound examination demonstrates inter-reader variability in qualitative steatosis assessment [12, 14–16]. MASLD is prevalent in up to 75% of the diabetic population, yet the shared pathophysiology between the two diseases and the influence of hepatic steatosis on type 2 diabetes progression requires further elucidation [17]. Exploring hepatic steatosis in diabetic patients through CT may provide insight into this relationship. Moreover, other hepatic and splenic parameters, such as liver and spleen volumes, can automatically be measured on CT scans using machine learning [18]. While hepatomegaly is well-associated with diabetes, the connection between splenic enlargement and diabetes has primarily been studied in animal models [19]. There remains a lack of literature on the precise quantitative relationship between the volume of these organs and the presence of type 2 diabetes. We seek to translate these findings and observe the degree of hepatosplenomegaly in our human diabetes cohort.

Diabetes is also associated with central adiposity, which comprises of visceral adipose tissue (VAT) located between organs and subcutaneous adipose tissue (SAT) located peripherally under the skin. While VAT is a known independent risk factor in type 2 diabetes pathogenesis, the relationship between subcutaneous adipose tissue (SAT) and type 2 diabetes remains unclear [2, 20, 21]. Moreover, visceral-to-subcutaneous fat ratio (VSR), a measure of body fat distribution, is associated with multiple cardiometabolic risk factors, but its association with type 2 diabetes was more recently found and requires further investigation [22–24]. Since machine learning algorithms have previously quantified abdominal and organ adipose tissue on CT, their use can be extended to characterize abdominal fat distribution in diabetic patients noninvasively [25–27].

These quantitative imaging measurements, or image-derived phenotypes (IDPs), are calculated from radiological scans, allowing us to explore novel disease associations. As IDPs are automatically quantified and established at the time of the study, their use presents no additional risk to the patient and provides an opportunity to detect findings that could be missed by radiologists, especially given the large volume of studies they typically read. Using the electronic health record (EHR) allows for long-term retrospective analyses of larger and more diverse cohorts [28].

In this study, we leveraged body composition analysis to uncover imaging markers of metabolic dysfunction in the diabetic population. We utilized EHR data from participants enrolled in the Penn Medicine Biobank to identify patients with non-contrast abdominal CT scans and a type 2 diabetes diagnosis. Using machine learning methods, we automated the analysis of certain abdominal IDPs, including SHAD as a marker for hepatic steatosis, LV, SV, VAT, SAT, and VSR. We aim to quantify the difference in these IDPs between nondiabetics and type 2 diabetics, as well as explore the predictive value of hepatic steatosis, abdominal fat distribution, and hepatosplenic organ size with type 2 diabetes presence.

## Materials and Methods

### Institution

This study utilized data from the Penn Medicine BioBank, a biomedical database consisting of advanced imaging, biological samples, and other EHR data from over 250,000 patients within the University of Pennsylvania Health System, a multi-hospital network headquartered in Philadelphia, PA. All enrolled patients provided informed consent for researchers to access their EHR.

### Machine Learning Algorithm for Adipose Quantification

A well-defined deep learning method that our group previously developed was applied for the segmentation and quantification of liver fat, liver and splenic volume, and abdominal adipose tissue [12, 26, 29]. In short, abdominal and pelvic CT scans designated by the appropriate Current Procedural Terminology (CPT) codes were selected **(Supplementary Figure 1)**. Axial CT scans that had high-pass filter kernel resolutions or those with image slice thickness < 2 mm were excluded to eliminate noise. Imaging studies with < 10 slices were excluded to avoid incomplete studies. The applied deep learning method consists of three convolutional neural networks (CNNs) **(Figure 1)**: CNN_1_ identified non-contrast CTs, as the contrast bolus affects peak splenic and hepatic attenuation in a time-dependent manner [12]. CNN_2_ defined certain axial slices as the abdominal compartment boundaries between the inferior thoracic cavity and L5 vertebrae. Lastly, fully automated segmentation of the liver (CNN_3A_), spleen (CNN_3B_), and delineation of SAT and VAT were conducted (CNN_3C_). Liver, spleen, SAT, and VAT cross-sectional areas were measured on each axial slice, with their respective total volumes computed as the sum across slices within the abdominal compartment. Liver and spleen mean attenuation values (LMA & SMA, respectively) were determined from the imaging scans to calculate SHAD, spleen attenuation minus liver attenuation. A greater or more positive value for SHAD corresponds with increased liver adiposity. We delineate hepatic steatosis through either a liver mean attenuation (LMA) < 40 Hounsfield units (HU) or a SHAD ≥ −1 HU. This value for SHAD has previously been found to represent the cutoff for mild hepatic steatosis, as the typical SHAD cutoff of 10 HU is actually more concordant with the cutoff for moderate-severe hepatic steatosis [14, 30]. Liver volume and spleen volume (LV and SV, respectively) were calculated by summation of the organs’ area on each axial slice.

**Figure 1:**
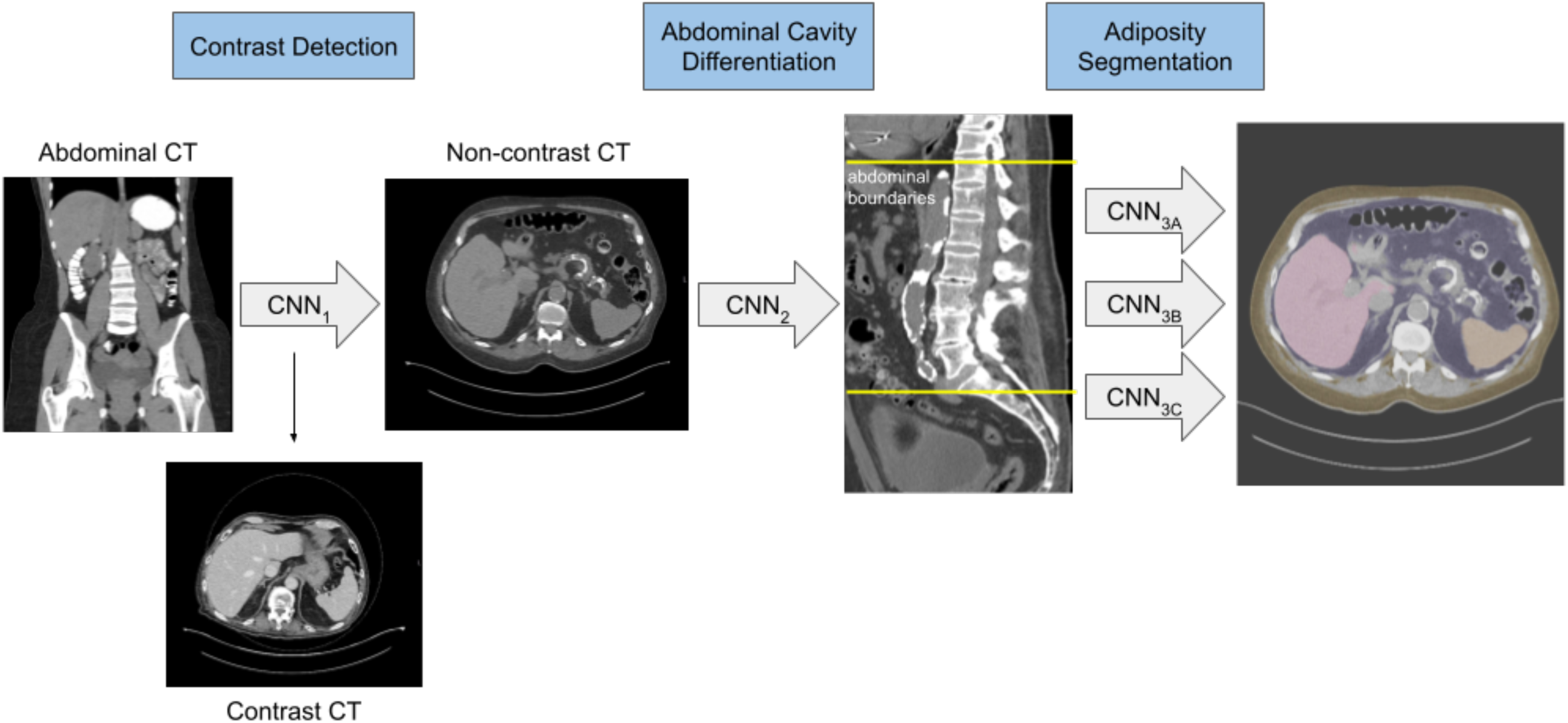
Deep learning method for the segmentation and quantification of hepatic and abdominal adiposity. CNN_1_ identified non-contrast from contrast CT scans. CNN_2_ delineated the boundaries of the abdominal compartment. The third CNN simultaneously segmented the liver (CNN_3A_) (in pink), spleen (CNN_3B_) (in orange), and characterized VAT (in purple) from SAT (in yellow) (CNN_3C_). Hepatic steatosis was diagnosed if SHAD was greater than −1 HU or LMA was less than 40 HU. CNN = convolutional neural network, VAT = visceral adipose tissue area, SAT = subcutaneous adipose tissue area, LMA = liver mean attenuation, SHAD = spleen hepatic attenuation difference, HU = Hounsfield unit.

### Study Cohort Imaging Data

9149 PMBB patients had abdominal CT scans that yielded IDP values for hepatic and abdominal adiposity [12, 30]. To associate a single SHAD value to each patient, we first selected the image series within each CT examination that had the SHAD value closest to the median SHAD of that entire exam. Of note, a patient can have multiple CT exams on different dates, and each CT exam has multiple series. Then, we grouped the data by PMBB identification number, which identified each unique patient, and selected the study that had the greatest SHAD value per patient. Patients with SHAD values within −30 HU and 30 HU were included. Certain inclusion and exclusion criteria were applied to obtain the patient population of interest **(Supplementary Figure 2)**. In brief, missing and extraneous IDPs were removed. Patients with missing age values were removed. We used mapped phecodes, which are disease phenotypes derived from the International Classification of Diseases, Ninth Revision (ICD-9) (https://www.phewascatalog.org/phecodes) to apply exclusionary criteria for certain diseases **(Supplementary Figure 3)** [31]. Those without diabetes and those with type 2 diabetes were included in the analysis. Patients with missing demographic information (race, BMI, sex) were excluded. VSR was calculated by dividing VAT by SAT. Patients without VSR or outlier values were excluded. Patients with alcohol use disorder, alcohol-related liver disease, end-stage liver disease and viral hepatitis phecodes were excluded to remove potential confounders. The final cohort comprised 5004 patients.

### Statistical Analysis

All statistical tests were done using R (R Core Team, version 4.3.1; Foundation for Statistical Computing, Vienna, Austria). Patients with at least one occurrence of the type 2 diabetes phecode were considered positive cases, whereas those with no occurrence of the general diabetes phecode were considered negative cases. Demographic information was collected. Moreover, IDPs from abdominal CT, such as LMA, SMA, SHAD, VAT, SAT, and VSR, were determined. Hepatic steatosis prevalence in each cohort was calculated. Differences in categorical variables (sex and race) between cohorts were determined using the chi-square test of independence. Differences in continuous variables (age, BMI, LMA, SHAD, VAT, SAT, VSR, HS prevalence) were determined using a Wilcoxon rank-sum test. Boxplots were created to compare IDP values with diabetes presence stratified by sex. Multivariate regression analyses were performed for the presence of type 2 diabetes, looking independently at IDPs (SHAD, LV, SV, VAT, SAT and VSR). The model shown below (Model 1) represents the six iterations of the regression analyses performed with each IDP, controlling for sex, age, BMI and race. Odds ratios (OR) were calculated from the multivariate analyses, including the 95% confidence interval. P-values were adjusted with Benjamini-Hochberg false discovery rate (BH-FDR) correction to reduce type 1 error with the statistical significance threshold of p<0.05.

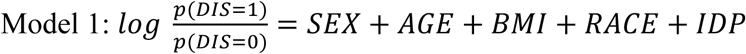

## Results

### Patient Cohort

Patients were dichotomized by the presence of the type 2 diabetes phecode **(Figure 2)**. Patients with type 2 diabetes were on average almost three years older than those without diabetes (63 years vs. 60 years, p = 6.59×10^-13^). The sex distribution in diabetic patients skewed more male (55.0% male) than non-diabetic patients (49.4% male, p = 1.32×10^-4^). The percentage of Black patients was greater for the type 2 diabetes group than the nondiabetic counterpart (37.4% vs. 20.0%, p < 2.2×10^-16^), while the percentage of White patients was lower for the type 2 diabetes group (55.0% vs. 73.9%). Patients with type 2 diabetes had a higher BMI than patients without diabetes (31.0kg/m^2^ vs. 27.2 kg/m^2^, p < 2.2×10^-16^).

**Figure 2:**
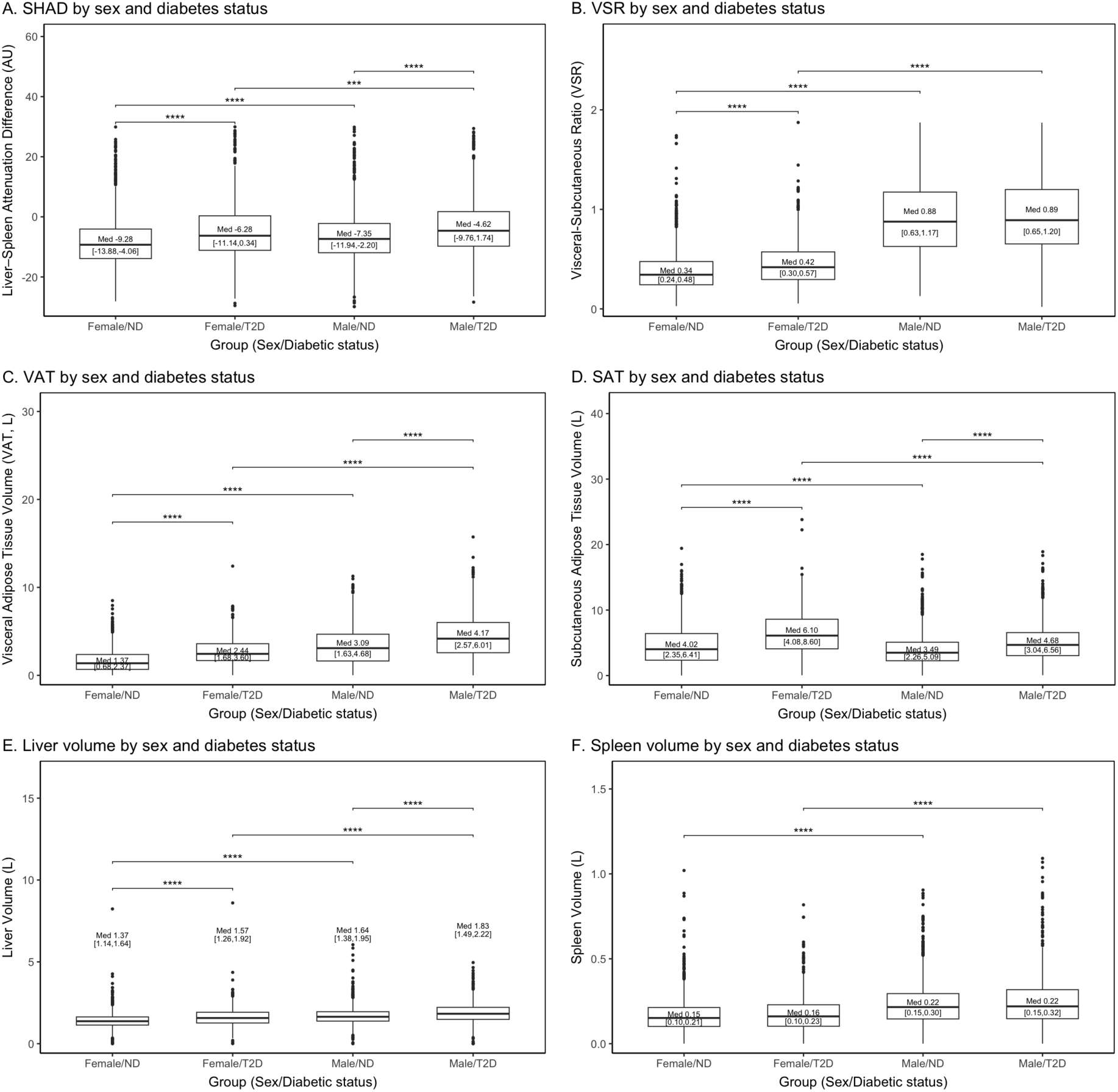
Box-and-whisker plots of IDPs in diabetic and nondiabetic patient cohorts categorized by sex. Boxplots show the distribution of image-derived phenotypes (IDPs) including liver–spleen attenuation difference (SHAD), visceral–subcutaneous fat ratio (VSR), visceral adipose tissue (VAT), subcutaneous adipose tissue (SAT), liver volume (LV), and spleen volume (SV) across four groups: Female/ND (non-diabetic), Female/T2D (type 2 diabetes), Male/ND, and Male/T2D. Median values are displayed above while IQR values are displayed below. Pairwise differences between groups were assessed using the Wilcoxon rank-sum test with Benjamini–Hochberg false discovery rate (BH-FDR) correction for multiple testing. Only significant pairwise comparisons are shown. Statistical significance is denoted by asterisks as follows: * *p* < 0.05; ** *p* < 0.01; *** *p* < 0.001; **** *p* < 0.0001.

Hepatic and abdominal IDPs were quantified stratified based on type 2 diabetes diagnosis **(Figure 3)**. Both SV (0.19 L vs. 0.18 L, p = 1.36×10^-3^) and LV (1.70 L vs. 1.50 L, p < 2.2×10^-16^) were larger in the type 2 diabetes patient cohort than in the non-diabetes cohort. LMA for diabetics was lower than for non-diabetics [47.6 HU vs. 52.4 HU, p < 2.2×10^-16^]. SHAD was greater in diabetics (–5.41 HU vs. –8.41 HU, p < 2.2×10^-16^). The presence of hepatic steatosis was greater in the diabetic cohort (38.4% vs. 21.4%, p < 2.2×10^-16^). The VAT volume (3.30 L vs. 2.06 L, p < 2.2×10^-16^) and VAT area (187.15 cm^2^ vs. 127.70 cm^2^, p < 2.2×10^-16^), as well as SAT volume (5.28 L vs. 3.72 L, p < 2.2×10^-16^) and SAT area (294.36 cm^2^ vs. 219.62 cm^2^, p < 2.2×10^-16^) were greater in diabetic patients. VSR was higher in patients with T2D (0.64 vs. 0.54, p = 5.86×10^-13^).

These trends generally held when stratifying by sex (**Figure 4)**, as women with diabetes showed significantly higher SHAD (–6.28 HU vs. –9.28 HU, p < 2.2×10^-16^), VSR (0.42 vs. 0.34, p < 2.2×10^-16^), VAT (2.44 L vs. 1.37 L, p < 2.2×10^-16^), SAT (6.10 L vs. 4.02 L, p < 2.2×10^-16^), and LV (1.57 L vs. 1.37 L, p < 2.2×10^-16^) than women without diabetes, and men with diabetes showed significantly higher SHAD (–4.62 HU vs. –7.35 HU, p = 9.38×10^-16^), VAT (4.17 L vs. 3.09 L, p < 2.2×10^-16^), SAT (4.68 L vs. 3.48 L, p < 2.2×10^-16^), and LV (1.83 L vs. 1.64 L, p = 1.81×10^-15^) than men without diabetes. However, VSR values between diabetic and nondiabetic males, as well as SV values between diabetic and nondiabetic males and females were nonsignificant. Between the sexes, median SHAD (−4.62 HU vs. −6.28 HU, p = 4.19×10^-4^), VSR (0.89 vs. 0.42, p < 2.2×10^-16^), VAT (4.17 L vs. 2.44 L, p < 2.2×10^-16^), LV (1.83 vs. 1.57 L, p < 2.2×10^-16^), and SV (0.22 L vs. 0.16 L, p < 2.2×10^-16^) were significantly higher amongst male diabetics, while SAT (6.10 L vs. 4.68 L, p = 9.82×10^-10^) volume was significantly higher in female diabetics.

### Multivariate Analysis

The multivariate analyses examined ORs of hepatic and abdominal IDPs with type 2 diabetes presence, controlling for age, sex, race, and BMI **(Figure 5)**. Please note that in this context, for continuous variables the odds ratio represents an increase in odds of diabetes presence per unit change for the quantity in question (i.e. increase per year of age or increase per unit of BMI). Age [ranging from OR 1.02 – 1.03, (p < 2.2×10^-16^)] and elevated BMI [ranging from OR 1.06 – 1.10, (p < 2.2×10^-16^)] were independently associated with the presence of type 2 diabetes. Males had either greater odds or no significantly greater odds of type 2 diabetes presence for different IDPs [ranging from OR 1.00 – 1.50, (lowest p = 1.88×10^-9^, greatest p = 0.99)]. Regarding race, Asians and Pacific Islanders were the most likely to have diabetes compared to the White cohort [ranging from OR 3.25 – 3.83, (greatest p = 3.57×10^-7^)], followed by Blacks [ranging from OR 2.38 – 2.92 (p < 2.2×10^-16^)]. Latinos [ranging from OR 1.97 – 2.06, (greatest p = 4.67×10^-3^)] and Others [ranging from OR 1.53 – 1.60, (greatest p = 3.57×10^-7^)] also showed statistical significance compared to Whites for diabetes.

When examining the IDPs, all six were independently associated with diabetes: SHAD [OR 1.04, (p <2.2×10^-16^)]; LV [OR 1.59, (p = 1.73×10^-11^)]; SV [OR 3.95, (p = 4.69×10^-7^)]; VAT [OR 1.23, (p <2.2×10^-16^)]; SAT [OR 1.05, (2.59×10^-3^)]; and VSR [OR 2.27, (p = 1.80×10^-13^)].

Even after removing BMI from the multivariate analyses **(Supplementary Figure 4)**, all of the IDPs were associated with diabetes: SHAD [OR 1.04, (p <2.2×10^-16^)]; LV [OR 2.37, (p <2.2×10^-16^)]; SV [OR 10.80, (p <2.2×10^-16^)]; VAT [OR 1.37, (p <2.2×10^-16^)]; SAT [OR 1.19, (p <2.2×10^-16^)]; and VSR [OR 1.97, (p = 3.19×10^-10^)].

## Discussion

In this study, we leveraged artificial intelligence applied to imaging and clinical data from a tertiary-care academic and community health system biobank to characterize hepatic and abdominal adiposity patterns in patients with and without diabetes. Our findings demonstrate robust associations on CT scans between various metrics associated with MASLD-related features, including elevated hepatic steatosis, hepatosplenomegaly, and the overall distribution of abdominal adipose tissue, with the presence of type 2 diabetes. The observed relationships suggest that metabolic dysfunction linked to peripheral insulin resistance are associated with increased liver and spleen volumes, increased visceral adipose tissue, and decreased attenuation of the organs indicative of fatty infiltration [32, 33].

The diabetic cohort was older and more likely to be male, which was also present in our multivariate analyses, concordant with evidence that men tend to have a greater prevalence of diabetes than women and that the disease tends to affect older individuals [1, 20, 24]. As type 2 diabetes can be considered a “lifestyle disease,” aging allows for disease state progression from glucose intolerance to actual type 2 diabetes [34]. Moreover, sex differences can contribute to this discrepancy between males and females, as estrogen protects against type 2 diabetes [35].

VAT is an alternative predictor of type 2 diabetes from BMI, and our findings suggest that VAT and VSR have a stronger association with type 2 diabetes presence than BMI [36, 37]. Because BMI is a flawed metric for obesity and can be affected by muscle mass [38, 39], VSR and VAT look at the distribution of abdominal fat and its relationship with other fatty compartments in the body to give a more complete understanding of how obesity contributes to type 2 diabetes [23, 24, 40, 41]. VSR has been shown to have prognostic value for type 2 diabetes, and thus our study highlights the growing importance of VSR as it relates to diabetes and metabolic disorders [22, 42]. Even after removing BMI from our regression, VSR still has an independent association with diabetes, suggesting that VSR captures a unique aspect of the etiology of type 2 diabetes. SAT was modestly associated with diabetes presence in our analysis (OR = 1.05), denoting that a 1 L increase in SAT volume is associated with a 5% increase in odds of a patient having diabetes. When interpreted in the context of its measurement scale and compared to other IDPs with the same units – such as VAT, LV and SV – SAT is less clinically relevant to diabetes. This aligns with previous findings suggesting that SAT’s relationship with diabetes is nuanced, with some evidence indicating that the specific location of abdominal SAT may even provide some protective effect against development of the disease [40, 43, 44]. Further research is necessary to comprehend the complex relationship between SAT and diabetes pathogenesis.

Hepatic steatosis is a predictor of type 2 diabetes, as these two phenotypes are correlated with each other [11, 17]. MASLD can contribute to the pathogenesis of type 2 diabetes through hepatic insulin resistance, which prevents hepatocytes from taking up glucose from the bloodstream [45]. While a SHAD of 10 HU is widely used by radiologists as the cutoff for moderate-to-severe hepatic steatosis, −1 HU has been histologically proven to coincide with the cutoff of at least mild steatotic liver disease [12, 14, 30]. By redefining the parameters of hepatic steatosis to make it more sensitive, we can capture a larger proportion of patients with milder hepatic steatosis.

One of this study’s strengths lies in the racial diversity of the PMBB patient population. Also, the biobank data is taken from patients who have diseases, whereas the UK Biobank consists mainly of healthy participants [46]. However, the biobank’s patient data is skewed to the geographic and demographic characteristics of southeastern Pennsylvania and New Jersey [47], although it does include patients from multiple hospitals and clinics in urban, suburban, and rural settings. A limitation of this study was the use of one CT scan to be representative of the patient. Although we could define associations between adiposity and diabetes, we were unable to determine a causal relationship of these IDPs to the onset of diabetes. Nevertheless, this study provided important insights to the relationship between fatty distribution and diabetes, guiding the design of future studies. Lastly, the use of phecodes to infer certain conditions presented some challenges since phecodes can imperfectly distinguish or group certain phenotypes [48].

In future research, we hope to further elucidate the relationship between diabetes incidence, hepatic steatosis, and abdominal fat distribution. A Korean study determined the VSR cutoff for men and women for predicting the incidence of type 2 diabetes, but no study has been done yet in America, which is more heterogeneous [24]. VSR holds promise as a metric to more accurately describe abdominal obesity and adiposity, and we hope to see it calculated and employed within clinical practice soon. Moreover, incorporation of the nutritional data of this patient population can provide insight into how diet and dietary modifications influence abdominal adiposity and disease etiology [49]. Current research is underway to explore how diet can cause changes in imaging before clinical manifestations occur.

Overall, hepatic steatosis, obesity, and diabetes are interlinked findings, and their early presentations potentially can serve as harbingers for more severe, even life-threatening disease. By applying artificial intelligence and using EHR data to characterize the relationship of IDPs with diabetes, we have confirmed and found novel associations that serve as a high-level overview to encourage future inquiry into how adiposity is related to diabetes mellitus. This study presents an accurate method for using machine learning to capture image-derived phenotypes in a diabetic population (or any other clinical population in the future) to catalyze translational science.

## Data Availability

The data supporting the findings of this study were obtained from the Penn Medicine BioBank and include human imaging and electronic health record data. Due to patient privacy considerations and institutional restrictions, these data are not publicly available. De-identified data may be made available to qualified researchers upon reasonable request and subject to approval by the Penn Medicine BioBank and relevant institutional review boards. Requests for data access should be directed to the corresponding author.

## Acknowledgements

We acknowledge the Penn Medicine BioBank (PMBB) for providing data and thank the patient-participants of Penn Medicine who consented to participate in this research program. The Institutional Review Board of the University of Pennsylvania approved the PMBB (IRB 808346, IRB 813913, IRB 817977).

## Author Contributions

R.T. - Conceptualization, Methodology, Validation, Formal Analysis, Investigation, Data Curation, Writing - Original Draft, Writing - Review and Editing, Visualization

P.R. - Conceptualization, Methodology, Software, Validation, Formal Analysis, Investigation, Data Curation, Writing - Review and Editing, Visualization

M.H. - Conceptualization, Methodology, Formal Analysis, Investigation, Data Curation

E.T. - Conceptualization, Methodology, Software, Formal Analysis, Investigation, Data Curation

S.S. - Methodology, Validation, Investigation

A.B. (Bhattaru) - Methodology, Validation, Investigation

M.M - Methodology, Software, Validation, Formal Analysis, Investigation, Data Curation

J.T.D. - Methodology, Software, Validation, Formal Analysis, Investigation, Data Curation

J.G. - Methodology, Resources, Data Curation

C.K. - Methodology, Validation, Investigation

D.J.R. - Resources, Data Curation

A.B. (Borthakur) - Methodology, Validation, Investigation

W.R.W. - Conceptualization, Methodology, Investigation, Resources, Data Curation, Writing - Review and Editing, Supervision

H.S. - Conceptualization, Methodology, Investigation, Resources, Data Curation, Writing - Review and Editing, Visualization, Supervision

## Competing Interests Statement

The authors declare no competing interests.

## Funding

W.R.W. is supported by NIH grants R01HL171709, R01HL169378, P41EB029460, R21EB036734, OT2OD038048. H.S. was supported by an RSNA Scholar grant through the Radiological Society of North America and was supported in part by the Institute for Translational Medicine and Therapeutics’ (ITMAT) Transdisciplinary Program in Translational Medicine and Therapeutics. The PMBB is supported by the Perelman School of Medicine at the University of Pennsylvania, a gift from the Smilow family, and the National Center for Advancing Translational Sciences of the National Institutes of Health under CTSA award number UL1TR001878. The content is solely the responsibility of the authors and does not represent the views of the Department of Veterans Affairs, the National Institutes of Health, or the United States Government. MTM received funding from the Sarnoff Cardiovascular Research Foundation.

## Prior Presentations

Parts of this paper was presented as an abstract at the 2023 American Roentgen Ray Society Annual Meeting in Honolulu, HI, USA. The citation is noted below:

Tran, R., Raghupathy, P., MacLean, M., Gee, J., Rader, D.J., Witschey, W.R., Sagreiya, H.: Hepatic steatosis in the diabetic population as assessed by using machine learning on CT scans. American Roentgen Ray Society Annual Meeting April 2023 Notes: Honolulu, HI.

---

**Table 1:** Patient Cohort Demographics.

| Diagnosis |  |  |  |
| --- | --- | --- | --- |
| Total (n=5004) | Nondiabetic (n=3278) | T2DM (n=1726) | P-value |
| **Age** (years) | 60 [49, 70] | 63 [55, 70] | $6.59 \times 10^{-13}$ |
| **Sex** (n(%)) | | | $1.32 \times 10^{-4}$ |
| Male | 1618 (49.4) | 950 (55.0) |  |
| Female | 1660 (50.6) | 776 (45.0) |  |
| **Race** (n(%)) | | | $< 2.2 \times 10^{-16}$ |
| White | 2421 (73.9) | 949 (55.0) |  |
| Black | 655 (20.0) | 645 (37.4) |  |
| Asian/PI | 52 (1.6) | 42 (2.4) |  |
| Latino | 58 (1.8) | 35 (2.0) |  |
| Other | 92 (2.8) | 55 (3.2) |  |
| **BMI** (kg/m<sup>2</sup>) | 27.2 [23.8, 31.5] | 31.0 [27.1, 36.5] | $< 2.2 \times 10^{-16}$ |

The demographic characteristics of included patients from the Penn Medicine BioBank. Age and sex are arranged as median [Q1, Q3]. Sex and race are organized as sample size (percentage). P-values for continuous variables (age, BMI) were calculated using a Wilcoxon rank-sum test and for categorical variables (sex, race) using a chi square test of independence to determine statistically significant differences between the “Nondiabetic” and “T2DM” groups. T2DM = type 2 diabetes mellitus, BMI = body mass index, PI = Pacific Islander.

**Table 2:** Imaging Derived Phenotype (IDP) Values Using CTs from the Patient Cohort.

| Diagnosis |  |  |  |
| --- | --- | --- | --- |
| Total (n=5004) | Nondiabetic (n= 3278) | T2DM (n= 1726) | P-value |
| Hepatic IDP Values |  |  |  |
| LMA (HU) | 52.4 [46.0, 57.6] | 47.6 [40.1, 53.9] | $<2.2 \times 10^{-16}$ |
| SMA (HU) | 44.2 [39.4, 48.0] | 42.2 [36.9, 47.5] | $2.07 \times 10^{-13}$ |
| SHAD (HU) | -8.41 [-13.0, -3.07] | -5.41 [-10.7, 1.16] | $<2.2 \times 10^{-16}$ |
| HS Prevalence (n(%)) | 702 (21.4) | 663 (38.4) | $<2.2 \times 10^{-16}$ |
| LV (L) | 1.50 [1.24, 1.82] | 1.70 [1.38, 2.08] | $<2.2 \times 10^{-16}$ |
| SV (L) | 0.18 [0.12, 0.26] | 0.19 [0.12, 0.28] | $1.36 \times 10^{-3}$ |
| Abdominal IDP Values |  |  |  |
| VAT volume (L) | 2.06 [0.96, 3.54] | 3.30 [2.01, 4.93] | $<2.2 \times 10^{-16}$ |
| SAT volume (L) | 3.72 [2.31, 5.61] | 5.28 [3.37, 7.70] | $<2.2 \times 10^{-16}$ |
| VAT area (cm<sup>2</sup>) | 127.70 [69.1, 202.49] | 187.15 [124.66, 265.89] | $<2.2 \times 10^{-16}$ |
| SAT area (cm<sup>2</sup>) | 219.62 [149.91, 202.19] | 294.36 [202.19, 424.62] | $<2.2 \times 10^{-16}$ |
| VSR | 0.54 [0.40, 0.98] | 0.64 [0.33, 0.90] | $5.86 \times 10^{-13}$ |

LV, SV, SMA, LMA, SHAD, VAT, SAT, and VSR are arranged as median [Q1, Q3]. HS prevalence for each group is formatted as sample size (percentage), where SHAD ≥ -1HU or LMA $<$ 40HU is classified as a HS diagnosis. P-values were calculated using a Wilcoxon rank-sum test to determine statistically significant differences between the “Nondiabetic” and “T2DM” groups and were corrected using Benjamini–Hochberg false discovery rate (BH-FDR) correction for multiple testing. T2DM = type 2 diabetes mellitus, LV = liver volume, SV = spleen volume, SMA = spleen mean attenuation value, LMA = liver mean attenuation value, SHAD = spleen-hepatic attenuation difference, HS = hepatic steatosis, VAT = visceral adipose tissue, SAT = subcutaneous adipose tissue, VSR = visceral-to-subcutaneous fat volume ratio, SHAD = spleen hepatic attenuation difference.

**Table 3:**
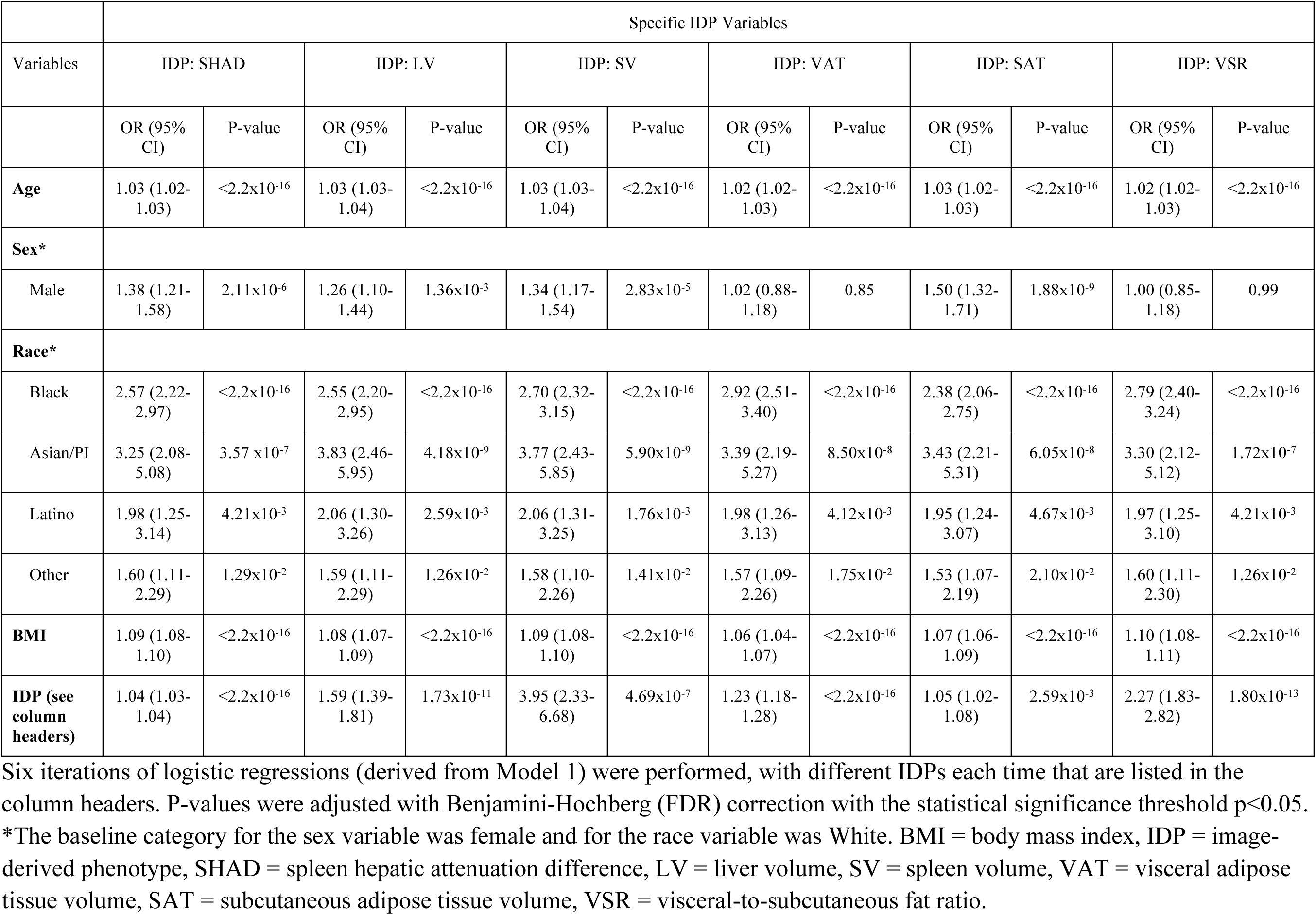
Odds ratio model of type 2 diabetes presence against abdominal adipose imaging phenotype values and demographic characteristics.

## Notes

### Competing Interest Statement

The authors have declared no competing interest.

### Author Declarations

IRB of the University of Pennsylvania gave ethical approval for this work

