## Supplementary Materials for "Hepatic and abdominal adiposity in type 2 diabetes as assessed with machine learning on CT scans"

Supplementary Figure 1: CPT codes

| CPT Code | Description |
|---|---|
| 74176 | CT abdomen and pelvis without contrast |
| 74178 | CT abdomen and pelvis with and without contrast |
| 74150 | CT abdomen without contrast |
| 74170 | CT abdomen with and without contrast |
| 71250 | non-contrast chest CT (diagnostic) |
| 71270 | non-contrast chest CT, with follow up CT scans (diagnostic) |

CPT codes used to identify computed tomography scans that had views of the abdomen. CPT = Current Procedural Terminology.

Supplementary Figure 2: Patient Flowchart

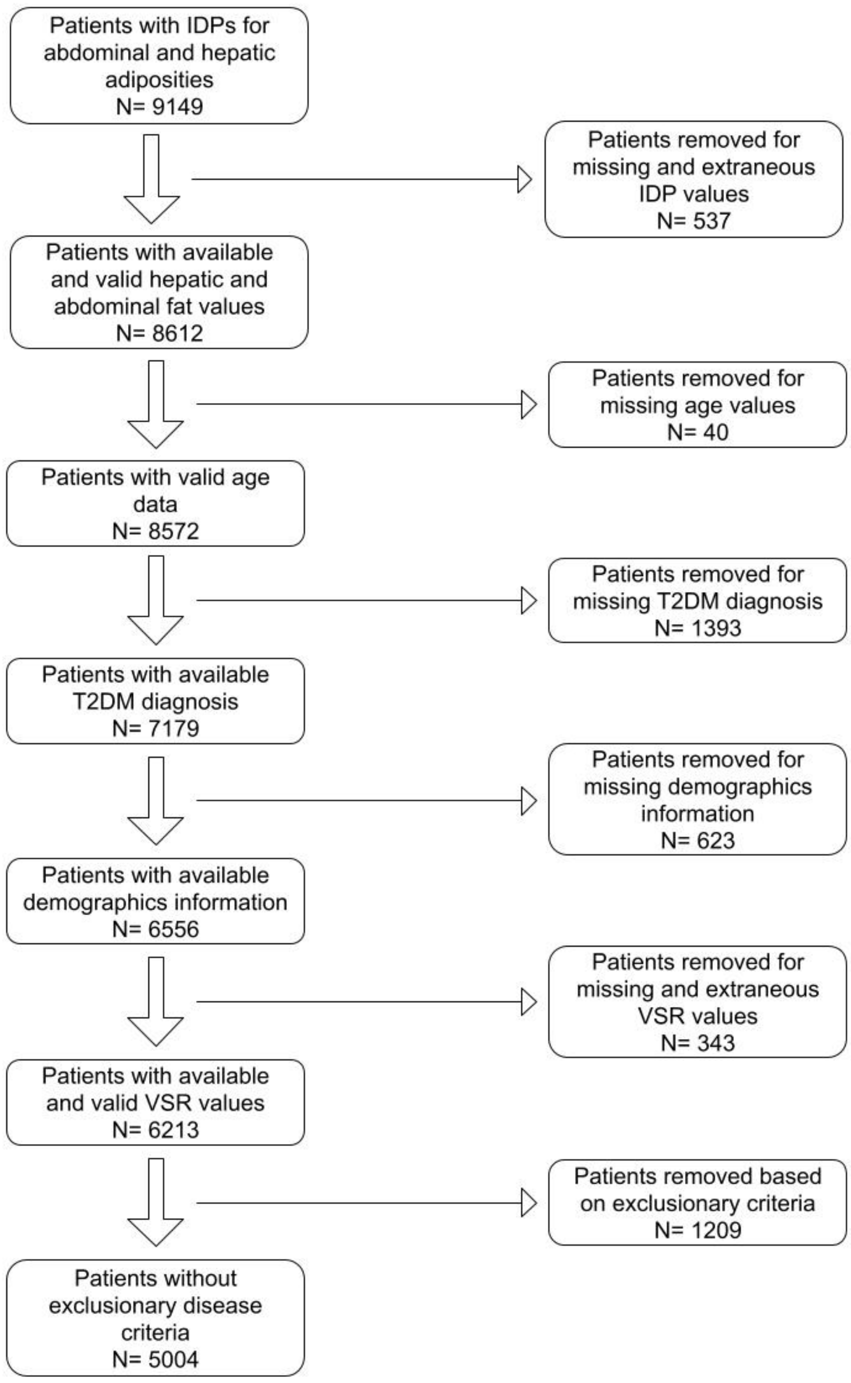


Number of patients in PMBB with available quantified image derived phenotypes related to hepatic and abdominal fat. Patients with missing demographics (Race, BMI, Sex, MRN), as well as patients with phecodes for exclusionary criteria (alcohol use disorder, hepatitis, and end-stage liver disease), were excluded from the final analysis. BMI = body mass index, T2DM = type 2 diabetes mellitus.

Supplementary Figure 3: ICD-9 codes and associated phecodes

| Phecode | ICD-9 Mapped Codes |
| --- | --- |
| Alcohol Use Disorders (317, 317.1, 317.11) | 291, 291.0-291.9, 303, 305.0, 305.00-305.03, 357.5, 535.3, 571.0-571.3, 790.3, 980.0 |
| Liver abscess and sequelae of chronic liver disease (571.8) | 572, 572.0-572.8 |
| Hepatitis (070, 070.1-070.4, 070.9) | 070. 573.1, 573.2, 571.3, 571.4 |
| Type 2 Diabetes Mellitus (250.2) | 250.00, 250.02, 250.10, 250.12, 250.20, 250.22, 250.30, 250.32, 250.40, 250.42, 250.50, 250.52, 250.60, 250.62, 250.70, 250.72, 250.80, 250.82, 250.90, 250.92 |

Phecodes are listed as phecode description (phecode number). Phecodes are mapped from ICD-9 (https://phewascatalog.org/phecodes). ICD-9 = International Classification of Diseases, Ninth Revision.

Supplementary Figure 4: Odds ratio model of type 2 diabetes presence against abdominal adipose imaging phenotype values and demographic characteristics without BMI

| | Specific IDP Variables | | | | | | | | | | | |
|---|---|---|---|---|---|---|---|---|---|---|---|---|
| Variables | IDP: SHAD | | IDP: LV | | IDP: SV | | IDP: VAT | | IDP: SAT | | IDP: VSR | |
| | OR (95% CI) | p-value | OR (95% CI) | p-value | OR (95% CI) | p-value | OR (95% CI) | p-value | OR (95% CI) | p-value | OR (95% CI) | p-value |
| **Age** | 1.02 (1.02-1.03) | $<2.2x10^{-16}$ | 1.03 (1.02-1.03) | $<2.2x10^{-16}$ | 1.03 (1.02-1.03) | $<2.2x10^{-16}$ | 1.02 (1.01-1.02) | $3.33x10^{-14}$ | 1.03 (1.02-1.03) | $<2.2x10^{-16}$ | 1.02 (1.01-1.02) | $2.81x10^{-12}$ |
| **Sex*** | | | | | | | | | | | | |
| Male | 1.28 (1.13-1.45) | $2.00x10^{-4}$ | 1.05 (0.92-1.19) | 0.50 | 1.18 (1.03-1.34) | $1.49x10^{-2}$ | 0.81 (0.70-0.93) | $4.20x10^{-3}$ | 1.56 (1.37-1.77) | $3.34x10^{-11}$ | 1.00 (0.85-1.17) | 0.96 |
| **Race*** | | | | | | | | | | | | |
| Black | 3.07 (2.67-3.54) | $<2.2x10^{-16}$ | 3.05 (2.65-3.51) | $<2.2x10^{-16}$ | 3.47 (3.00-4.01) | $<2.2x10^{-16}$ | 3.51 (3.04-4.06) | $<2.2x10^{-16}$ | 2.49 (2.16-2.87) | $<2.2x10^{-16}$ | 3.28 (2.84-3.79) | $<2.2x10^{-16}$ |
| Asian/PI | 2.27 (1.48-3.50) | $2.88x10^{-4}$ | 3.25 (2.11-5.00) | $1.62x10^{-8}$ | 2.90 (1.89-4.45) | $1.76x10^{-6}$ | 2.87 (1.86-4.44) | $3.15x10^{-6}$ | 2.96 (1.92-4.57) | $1.62x10^{-6}$ | 2.28 (1.48-3.47) | $1.41x10^{-4}$ |
| Latino | 2.05 (1.31-3.17) | $2.09x10^{-3}$ | 2.21 (1.42-3.46) | $6.96x10^{-4}$ | 2.22 (1.43-3.44) | $5.59x10^{-4}$ | 2.03 (1.29-3.18) | $2.72x10^{-3}$ | 1.95 (1.24-3.06) | $4.44x10^{-3}$ | 2.06 (1.33-3.20) | $1.67x10^{-3}$ |
| Other | 1.61 (1.13-2.29) | $9.57x10^{-3}$ | 1.66 (1.16-2.36) | $6.27x10^{-3}$ | 1.63 (1.15-2.32) | $7.49x10^{-3}$ | 1.60 (1.11-2.30) | $1.33x10^{-2}$ | 1.51 (1.06-2.16) | $2.36x10^{-2}$ | 1.61 (1.14-2.29) | $8.41x10^{-3}$ |
| **IDP (see column headers)** | 1.04 (1.04-1.05) | $<2.2x10^{-16}$ | 2.37 (2.09-2.68) | $<2.2x10^{-16}$ | 10.80 (6.53-17.88) | $<2.2x10^{-16}$ | 1.37 (1.32-1.41) | $<2.2x10^{-16}$ | 1.19 (1.16-1.21) | $<2.2x10^{-16}$ | 1.97 (1.60-2.42) | $3.19x10^{-10}$ |

Six iterations of logistic regressions (derived from Model 1 without BMI) were performed, with different IDPs each time that are listed in the column headers. P-values were adjusted with Benjamini-Hochberg correction with the statistical significance threshold of $p<0.05$. *The baseline category for the sex variable was female and for race variable was White. IDP = image-derived phenotype, SHAD = spleen hepatic attenuation difference, LV = liver volume, SV = spleen volume, VAT = visceral adipose tissue volume, SAT = subcutaneous adipose tissue volume, VSR = visceral-to-subcutaneous fat ratio.
